# Transient loss and recovery of oral chemesthesis, taste and smell with COVID-19: a small case-control series

**DOI:** 10.1101/2023.03.27.23287763

**Authors:** Elisabeth M. Weir, Cara Exten, Richard C. Gerkin, Steven D. Munger, John E. Hayes

## Abstract

Anosmia is common with respiratory virus infections, but loss of taste or chemesthesis is rare. Reports of true taste loss with COVID-19 were viewed skeptically until confirmed by multiple studies. Nasal menthol thresholds are elevated in some with prior COVID-19 infections, but data on oral chemesthesis are lacking. Many patients recover quickly, but precise timing and synchrony of recovery are unclear. Here, we collected broad sensory measures over 28 days, recruiting adults (18-45 years) who were COVID-19 positive or recently exposed (close contacts per U.S. CDC criteria at the time of the study) in the first half of 2021. Participants received nose clips, red commercial jellybeans (Sour Cherry and Cinnamon), and scratch-n-sniff cards (ScentCheckPro). Among COVID-19 cases who entered the study on or before Day 10 of infection, Gaussian Process Regression showed odor identification and odor intensity (two distinct measures of function) each declined relative to controls (close contacts who never developed COVID-19), but effects were larger for intensity than identification. To assess changes during early onset, we identified four COVID-19 cases who enrolled on or prior to Day 1 of their illness – this allowed for visualization of baseline ratings, loss, and recovery of function over time. Four controls were matched for age, gender, and race. Variables included sourness and sweetness (Sour Cherry jellybeans), oral burn (Cinnamon jellybeans), mean orthonasal intensity of four odors (ScentCheckPro), and perceived nasal blockage. Data were plotted over 28 days, creating panel plots for the eight cases and controls. Controls exhibited stable ratings over time. By contrast, COVID-19 cases showed sharp deviations over time. No single pattern of taste loss or recovery was apparent, implying different taste qualities might recover at different rates. Oral burn was transiently reduced for some before recovering quickly, suggesting acute loss may be missed in data collected after acute illness ends. Changes in odor intensity or odor identification were not explained by nasal blockage. Collectively, intensive daily testing shows orthonasal smell, oral chemesthesis and taste were each altered by acute COVID-19 infection, and this disruption was dyssynchronous for different modalities, with variable loss and recovery rates across modalities and individuals.

## 1. Introduction

The COVID-19 pandemic caused by the SARS-CoV-2 virus is one of the most devastating infectious disease outbreaks since the H1N1 avian flu of 1918 [1, 2]. By the end of 2021, roughly two years after the start of the pandemic, there were over 281 million cases of COVID-19 globally, resulting in over 5.4 million deaths [3]. Early in the pandemic, SARS-CoV-2 infection was associated with myriad symptoms, one of the most common being anosmia [4–7]. Meta-analyses of dozens of early studies suggested half to three-quarters of COVID-19 patients lost their sense of smell [8]. Further, smell loss was the most predictive symptom of COVID-19 [9] in the first several waves.

In contrast to other respiratory illnesses that cause acute anosmia – including those caused by rhinoviruses, influenza viruses, and common coronaviruses – both taste and chemesthesis function were reportedly lost in some people with COVID-19 [10–12]. Taste loss in the absence of smell loss is rare [13], and many individuals may mistakenly conflate the impaired flavor perception associated with anosmia with a true loss of taste function. However, one large crowd-sourced study reported ∼60% of COVID-19-positive individuals had impaired perception of specific taste qualities (i.e., sweet, salty, sour or bitter tastes), suggesting taste loss in these individuals is distinct from impaired flavor perception accompanying smell loss [10, 14]. The findings of that study and others based on self-reports (e.g., [15–17]), were confirmed by psychophysical tests of taste function, suggesting ∼47 to 64% of COVID-19 positive individuals experience taste loss [18, 19]. As a result, taste dysfunction (distinct from impaired flavor perception due to smell loss) is now also recognized as a common symptom of COVID-19 [20, 21].

Data on disruption of chemesthesis associated with COVID-19 remains quite limited. Consistent with many patient anecdotes that chili and ethanol burn were transiently depressed (e.g., [22]), studies relying on self-report suggested roughly half of individuals with COVID-19 experienced disruptions of chemesthesis [10]. In a small study of Italians with COVID-19-associated smell loss, 57% of patients had reported a severe impairment of nasal chemesthesis at initial diagnosis, but over 90% reported full recovery of chemesthesis six months later [23]. A Swedish study that used “olfactory” stimuli known to concomitantly activate the trigeminal system (e.g., vinegar, chopped garlic, vodka) provided evidence for impairment of nasal chemesthesis [24]. Thus, while self-report and clinical assessment both suggest COVID-19 may associate with acute impairment of chemesthesis in many individuals, the time course of loss and recovery is lacking, as is any assessment of the impact on oral chemesthesis. We attempt to fill these knowledge gaps here.

In approximately 85% of COVID-19 cases where chemosensation (smell, taste and/or chemesthesis) has been affected, recovery of chemosensory function is typically seen within ∼6 weeks [6, 25, 26]. Unfortunately, some patients do not report appreciable recovery after many months [13, 26–30]. However, without daily chemosensory testing, the precise timing of recovery remains unknown [6, 25, 31]. Because patient anecdotes suggest the timing of recovery of different chemical senses may be dyssynchronous [26], we assessed smell, taste, and chemesthesis function acutely and transiently over 28 days by collecting longitudinal data using commercially available chemosensory stimuli in a cohort of patients diagnosed with COVID-19 and control participants without COVID-19 in early 2021. Here, we present a small case-control series using temporally intensive data collection from remote daily testing. We focus on a handful of COVID-19 cases that allow for visualization of loss and recovery of chemosensory function as well as baseline ratings obtained prior to the onset of illness.

## 2. Methods

### 2.1 Study design and recruitment

This prospective study investigated COVID-19-related chemosensory dysfunction in a community-derived sample of 18- to 45-year-old adults recruited on and around the campus of a large public university in rural central Pennsylvania (i.e., the area surrounding State Colleg<u>e</u>, PA). Enrollment began in February, 2021, on a rolling basis using geotargeted ads on social media, and ended in May, 2021. Potentially interested individuals were asked to contact a study team member (author EMW) via email if they believed they were qualified for the study, who then emailed them a link to a brief screening questionnaire. The screening questionnaire asked questions about demographics, prior diagnosis of COVID-19, contact with a COVID-19-positive (COVID- 19+) individual, and any recent symptoms of COVID-19. Contact with a COVID-19+ individual was defined by the Centers for Disease Control (CDC) screening criteria in use at the time of enrollment (specifically, 15 or more minutes within 6 feet of a confirmed case of COVID-19). Due to the fluidity of the COVID-19 pandemic in early 2021, however, a strict timeline was not rigidly enforced and enrollment occurred on a case-by-case basis. As vaccines were not available to non-health care workers at study initiation in February, 2021, but became more widely available during the enrollment window, we added a short retroactive questionnaire to the end of the study to gather self-reported information on vaccination status and date. No attempt was made to confirm these reports against medical records.

Participants with the following conditions were excluded: not diagnosed with COVID-19 or not a close contact of a COVID-19+ individual, pregnant, food allergies (or another reason they could not consume commercial jelly beans), prior history of a disease of the central nervous system (including Alzheimer’s disease, multiple sclerosis, Parkinson’s disease, Huntington’s disease, brain tumor), nasal obstruction (tumor/polyps), a history of nasal surgery, history of a severe head injury/concussion, history of chronic sinus infections, history of radiation therapy to the head or neck (ever), recent chemotherapy (within the last year), a prior diagnosis of smell or taste loss, diabetes, history of lung/pulmonary disease or neurological disease, were unwilling to create a PayPal account for compensation if they did not already have one, or were below 18 years or above 45 years of age.

Data were collected using REDCap, a secure data capture platform for clinical research [32, 33] on a server hosted and maintained by the Penn State College of Medicine in Hershey, PA. The study was performed in compliance with the principles of the Declaration of Helsinki, informed consent was obtained electronically, and the specific protocol was approved by the Institutional Review Board at Penn State (STUDY00016377). The subject identification numbers referenced below were known only to our research staff and were not known to the participants or other individuals.

### 2.2 Chemosensory stimuli and assessment

The longitudinal design consisted of brief daily assessment every day for 28 days, followed by four additional follow-up sessions every 2 weeks, for a total of 32 sessions over a 12-week period. This report focuses on data from the first 28 days of testing. In each daily session, participants were asked to complete questions on COVID-19 status or symptoms that had changed since the last session, as well as self-administered psychophysical smell and taste tests. They were instructed to minimize any distracting smells or odors before any sensory testing and were also asked not to eat or drink (anything other than water) or smoke for at least 30 minutes prior to testing.

Upon enrollment, the first author arranged contactless delivery of all research materials in a large plastic zip-top bag. Specifically, participants were given 32 ScentCheckPro cards (Item #098515, Lot #0821) from Taylor Corp (North Mankato, MN). Each ScentCheckPro card consisted of 4 microencapsulated scents in a scratch-n- sniff format, with one scent located near each corner of the postcard sized card. The four scents on a given card were some combination of the following: coconut, grape, coffee, lemon, bubble gum, popcorn, pine, cinnamon, flowers, banana, or none of these.

Participants were also given 36 lidded plastic 2 oz souffle cups (Solo P200N; Lake Forest IL) labeled with blinding codes. Individual cups contained one of two kinds of red colored jellybeans (Jelly Belly, Fairfield, CA): either Sour Cherry (Lot #20200601) or Cinnamon (Lot #200731). Each cup contained three jellybeans of a single flavor. Jellybeans are a highly familiar confection in the United States that are made with sweeteners (sugar, and/or corn syrup), corn starch, confectioners glaze, added color and natural or artificial flavors. They have a shiny candy shell and a soft gel center, and come in many different colors and flavors including fruit or spice flavors. With the nose pinched closed, the Sour Cherry jellybean used here evoke both sweetness and sourness, while the Cinnamon jellybean elicits sweetness and a mild warming/burning sensation. Because of the glazed outer shell, jellybeans have little to no orthonasal smell, and both jellybeans had a similar red color, so there were no obvious cues of the specific flavor in each cup. Across days, jellybean presentation order was counterbalanced with pairwise randomization, so that a participant who was presented with Sour Cherry on Day 1 would get Cinnamon on Day 2, while the next participant would start with Cinnamon on Day 1 before receiving Sour Cherry on Day 2. This procedure was used to maximize the range of chemosensory stimuli used in the study (i.e., taste, smell, and chemesthesis) while minimizing participant burden on any given day of the study (i.e., a very brief test time to enhance compliance). Individual lidded cups were labeled with random 3-digit blinding codes, and these codes were programmed into REDCap prompts for each session to help ensure participants sampled the correct jellybean on the correct day. We also provided a disposable foam padded nose clip (A-M Systems; Sequim WA; Model #166500, Lot #189615) to allow ratings of oral sensation to be collected with occluded nostrils to minimize olfactory input.

Once a day, participants were asked to rate how blocked their nose was using a horizontal 0-100 visual analog scale (VAS) scale anchored with ‘Not blocked at all’ to ‘Completely blocked’. Participants were then asked to scratch the spot containing the encapsulated odorant on each postcard sized smell card with a coin or fingernail for 5-10 seconds before sniffing; they were then asked to bring the card one inch from their nose and sniff the odor. For each odor spot, participants were asked to first identify the odor they smelled from four multiple choice options presented in REDCap before rating the perceived intensity of the odor on a 0-100 VAS anchored with labels of ‘None’ to ‘Very intense’. This process was repeated for the four different odorants on a given card.

Next, participants were asked to pinch their nose closed using the provided nose clip and put all three jellybeans from the cup into their mouth. With their nose *pinched closed*, they were asked to chew the jellybeans slowly, and rate the perceived intensity of various qualities on five different horizontal 0-100 VAS scales labeled ‘None’ to ‘Very intense’. These qualities included: *sourness*, *sweetness*, *warming/burning*, *cherry flavor*, and *cinnamon flavor*. Participants were then asked to *unpinch* their nose and exhale (while still chewing the jellybeans) and rate the same five intensity scales with their nose *unpinched* (data not shown). All five scales were presented in both conditions (nose closed / nose open) to minimize any “dumping” artifacts [34]. To decrease daily test time and participant burden on days 1 through 28, participants assessed only one jellybean flavor per day (either Sour Cherry or Cinnamon, counterbalanced as described above). In four follow up sessions at weeks 6, 8, 10 and 12, a longer assessment was deemed reasonable, so participants were presented with two cups of jellybeans, one with each flavor. These data are not reported here.

### 2.3 Categorization of participants as COVID-19 cases versus controls

For the purposes of this study, a *COVID-19 contact* was defined as a participant who had been exposed to a COVID-19+ individual (e.g., 15 or more minutes within 6 feet of a confirmed case of COVID-19, per US CDC guidelines at the time), but never developed any symptoms or received a positive diagnosis in subsequent testing. Conversely, a *COVID-19 case* was defined as a participant who was either formally diagnosed with COVID-19 *or* began having symptoms while enrolled in the study following their recent exposure. By studying close contacts who later became cases while enrolled in our study, we were able to observe acute changes in chemosensation using controlled stimuli from the earliest days of their infection. When COVID-19 symptoms started prior to the participant receiving a positive COVID-19 diagnosis via clinical testing (typically a positive PCR test), an *estimated day of infection (Day 0)* was defined as the first day of symptoms. When a positive test preceded COVID-19 symptoms, the date of the positive test was used as *Day 0 of infection*. Data from controls were not centered on the day of infection, as these participants enrolled at their discretion. For these individuals, the *day of rating (Day 0)* is defined as the first day of their participation in the study.

### 2.4 Data analysis

Between February and May of 2021 (i.e., in the months prior to the Delta and Omicron waves in North America), a total of 55 participants were enrolled in the study. For this analysis, 39 participants with confirmed COVID-19 infection were identified as *COVID- 19 cases* (Figure 1). Of these 39, 15 participants were identified as having an active COVID-19 infection and enrolled in the study prior to or during the first 10 days of infection. Of these 15 participants, four COVID-19 cases entered the study on or before day 1 of their infection, allowing us to capture acute changes in their symptoms throughout their entire infectious period, including early onset of symptoms. As shown in Figure 1, three more cases enrolled on days 2-4 of infection, and nine enrolled on day 5-10 of their infection. An additional 24 participants who had enrolled were identified as COVID-19 cases, but only after their initial 10-days of infection had passed. Because we were not able to enroll these subjects early enough to capture potential changes in chemosensory function during acute illness, data from these subjects were not included in the present analyses. We plan to analyze and publish these data elsewhere to better understand recovery over time, but the focus of the current report is initial loss during early illness. Finally, in this analyses, 15 participants were identified as *controls* (i.e., they did not develop COVID-19 during the duration of the study).

**Figure 1:**
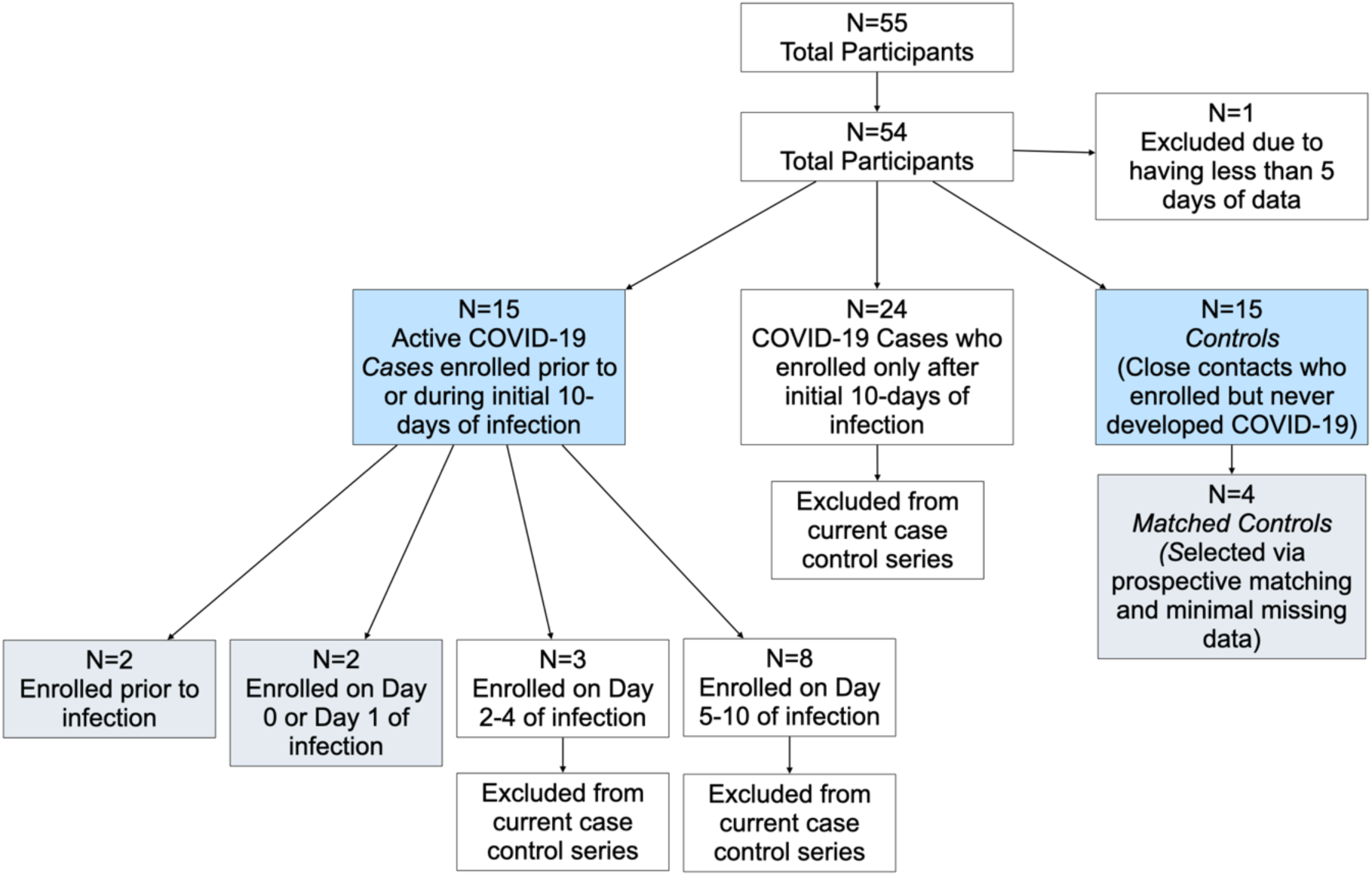
Flow diagram summarizing selection of cases and controls for this case series. Blue boxes indicate the 30 participants included in the Gaussian Process model regression, and gray boxes indicate the 8 individuals shown in the in-depth panel plots.

Here, we wanted to assess whether deviation of smell intensity in the cases differed from controls, so we identified the fifteen COVID-19 cases in our study who had enrolled on or before day 10 of either infection and tested if their average deviation in smell intensity over time differed from the 15 participants who were controls. To do this, a grand mean of smell intensity was calculated for the controls across all days and individuals. Deviation scores (deltas) for the 15 COVID-19 cases were then calculated by subtracting that individual’s ratings from the grand mean of the 15 controls, resulting in a deviation score for smell intensity. Similarly, for the controls, we subtracted each control’s *individual* rating from the *grand mean* of all controls, to get a deviation score for that rating relative to performance of the group. We used a Gaussian Process Regression model to analyze the deviation (delta) scores for *cases* and *controls* over time (both individually and as a group). A Gaussian Process model is a probabilistic unsupervised machine learning concept used for regressions in which the model makes predictions by utilizing prior knowledge about the smoothness of plausible time series and provides uncertainty measures for such predictions [35]. COVID-19 cases were normalized on Day 0 of infection and missing values were extrapolated via the Gaussian Process Regression model. Controls were normalized on Day 0 of rating. To be conservative and avoid overfitting sparse data, ratings of the two jellybean flavors were not modeled via Gaussian Process Regressions, as the counterbalancing of flavors across days meant only half as many data points were available for analysis. Analyses and data visualization were conducted using SAS software (Version 9.4), R using RStudio software (Version 2021.09.0), Python software (Version 3.9.10), or DataGraph version 4.7.1 (Visual Data Tools, Inc; Arlington TX).

Elsewhere, it has been suggested smell and taste changes occur within the first four days of disease onset [36], and the median incubation period for symptom onset is approximately five days [37]. We observed the same overall pattern within our data; as reported below, it became clear the confidence interval of cases did not include zero in the first week of infection. Given the unique opportunity to explore acute and early changes in chemosensation in a small number of participants who had been enrolled prior to acute illness, we also performed an in-depth analysis of these individuals, in hopes of better understanding the initial trajectory of changes in chemosensation.

### 2.4 Selection of participants for in-depth case control series

Here, we describe a very small case series restricted to four specific cases who enrolled in the study prior to or on Day 1 of infection (Figure 1). Studying these four participants in detail allowed us to capture changes in symptoms throughout their entire infectious period, including early onset of symptoms. More specifically, this prospective approach maximizes the potential to capture baseline ratings, loss, and recovery of chemosensory function that would not be possible with patients recruited only after they were ill. This is a distinct and unique feature of this case series, as most other studies have assessed chemosensory function multiple days into a participant’s isolation period, which does not allow for visualization of initial loss [30, 38–43]. For the other 12 participants who entered on or before Day 10 of infection (see Figure 1), we did not capture their initial loss, so they are not included in this analysis.

From the 15 *controls*, four were selected as *matched controls* for the four COVID-19 *cases*, based on age, gender, and race (Figure 1). To be considered for matching with a specific COVID-19 case, potential candidates were required to (a) provide data on a minimum of 80% of days, and (b) remain active for the full 28-day data collection period (to avoid bias from dropout over time). If more than one potential candidate met the age, gender, and race criteria to be included in a matched pair, the control used here was randomly selected. Matching was performed manually by an experienced epidemiologist (author CE). This process resulted in a final analysis of four *cases* and four *matched controls* (Figure 1; Table 1).

**Table 1:**
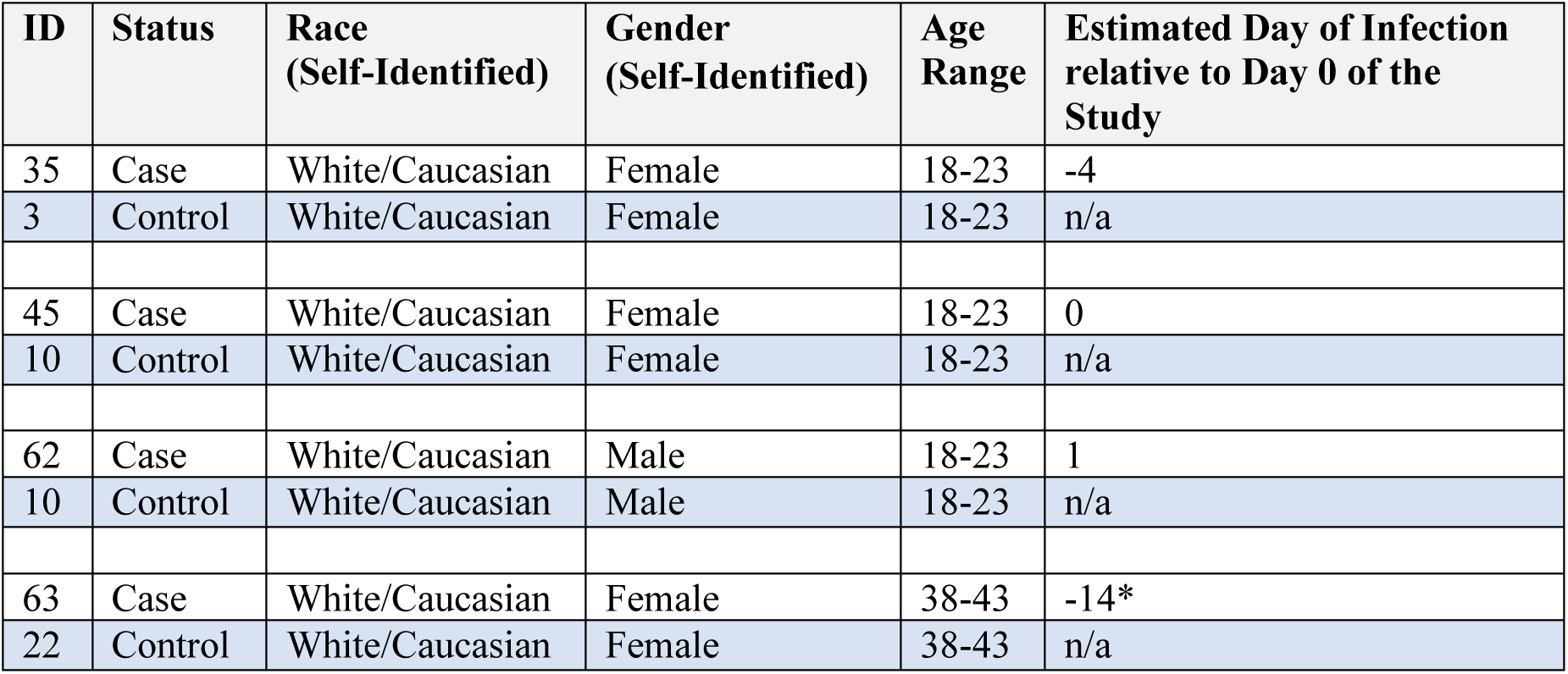
Participant demographics from COVID-19 case-control series identifying cases and matched controls.

For these four *cases* and their *matched controls*, we plotted six key variables related to smell, taste, and chemesthesis. Specific outcomes were selected *a priori* by two authors (EMW and JEH) as being the most salient and theoretically interesting variables. These were: perceived *nasal blockage,* mean orthonasal *smell intensity* of the four odorants on a given ScentCheckPro card*, sourness* and *sweetness* (from the Sour Cherry jellybeans), and oral *burn* (from the Cinnamon jellybeans). Also, we included *daily number correct* on the odor identification task. These seven variables were then plotted across all 28 days to create a series of panel plots for all eight individuals.

In summary, ratings were collected on 101 point VAS, and variables summarized here were: daily ratings of *nasal blockage*; a daily measure of orthonasal *smell intensity* derived from the mean of four scratch-n-sniff spots on a ScentCheckPro card for a given day; oral *burn* ratings collected every other day from a Cinnamon jellybean; and *sweetness* and *sourness* ratings collected every other day from a Sour Cherry jellybean.

## 3. Results and Discussion

### 3.1 Odor identification scores and ratings of orthonasal intensity from a commercial scratch-n-sniff card

For controls, the grand mean correct on the daily odor identification (OdorID) across the entire study period was 3.32 out of 4 possible (Figure 2, Supplemental Figures 1, 2). There was some evidence of a learning effect, as controls got slightly better at the task over time (see upward slope, Figure 2A and 2B). For OdorID, the delta score for cases deviates from zero, with a maximal dip occurring around days 5 to 8 (Figure 2A and 2B). Conversely, this dip was not observed in the controls, indicating that the drop in OdorID performance was limited to COVID-19 cases.

**Figure 2.**
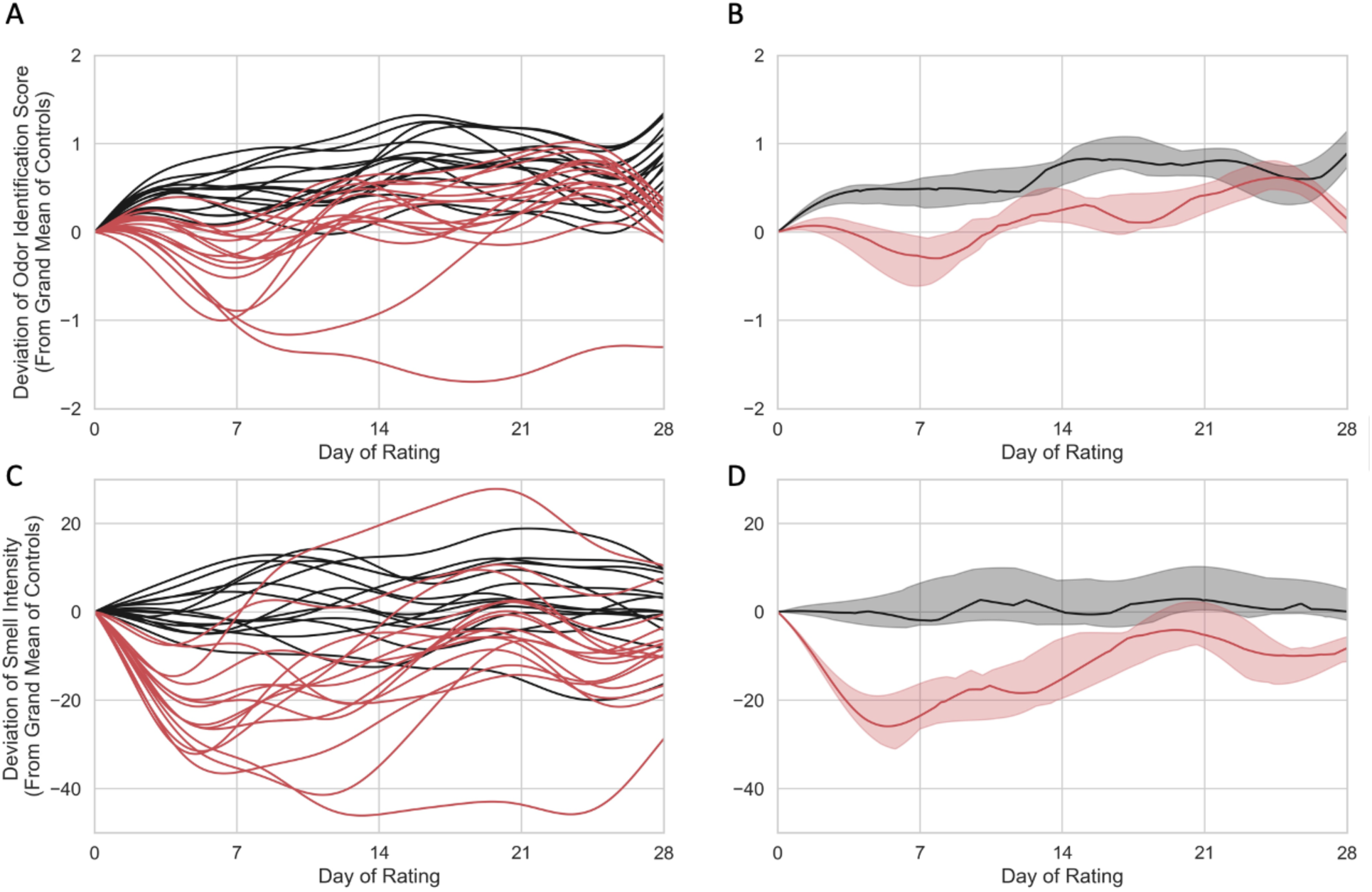
Individual (left) and group level (right) Gaussian Process Regression models for 15 controls (red) and 15 COVID-19 cases (black) who entered the study on or before day 10 of their infection. For cases, Day of Rating is centered on estimated day of infection. Odor identification scores (top) and odor intensity ratings (bottom) are shown as deviation scores calculated from a grand mean of controls across time. **(A)** Individual odor identification data in a Gaussian Process Regression model of deviation scores from 15 COVID-19 cases (red) and 15 controls (black). **(B)** Group level odor identification data in a Gaussian Process Regression model for COVID-19 cases (red) and controls (black). **(C)** Individual deviation scores for smell intensity ratings for 15 COVID-19 cases (red) and 15 controls. **(D)** Group level smell intensity data in a Gaussian Process Regression model for 15 COVID-19 cases (red) and 15 controls (black). In panels B and D, the solid line represents the group median, and the shaded region shows the interquartile range. Panels A and C show maximal loss roughly around day. Panel B shows some evidence of a learning effect for the OdorID task in the controls; no evidence of learning is seen for controls in the intensity task shown in panel D.

After performing the OdorID task for 4 days, the median number correct for controls increased by ∼0.5, and after 14 days (two weeks), the median number correct for controls increased by ∼0.7-0.8 above the grand mean; that is, there was nearly perfect performance on a 4 item OdorID task. Based on odor intensity ratings (discussed below), smell loss for cases appears to be maximal near Day 5 (see Figure 2C and 2D). Accordingly, we would also expect OdorID performance to be lowest on Day 5. However, this is just when the first increase due to the learning effect appears to occur (Figure 2A and 2B). If we assume cases (at least those who are not totally anosmic) show similar ability to learn as the controls, this offsetting bump upward would minimize the apparent dip in OdorID performance seen around Day 5. Thus, the COVID-19-associated drop in identification performance may appear smaller than it actually is due to a simultaneous offsetting increase in performance due to learning (or practice). Evidence of such learning is also seen in Supplemental Figures 1 and 2. Regarding suprathreshold odor intensity, the grand mean of perceived intensity ratings for controls across time was 59.47 on 0 to 100 VAS. The mean on Day 0 was used to calculate daily deviation scores for cases and controls (Figure 2C and 2D). Data were centered using Day 0 means so the delta could be more easily compared across the data set. For the cases (Figure 2C and 2D), the delta score of smell intensity clearly deviates from zero, and this difference was maximal in the first week of infection, as noted above. In sharp contrast, this pattern is not seen in the controls, with a score of zero falling inside the interquartile range across the entire study period (Figure 2D). This indicates there was a significant drop in smell intensity for cases but not controls for ratings of the scratch-n-sniff spots on commercial ScentCheckPro cards. In contrast with the learning seen for the OdorID data, we failed to observe any evidence of a learning effect for smell intensity ratings. Collectively, these data suggest OdorID tasks may be less sensitive to acute changes in smell with COVID-19, relative to odor intensity ratings, at least in repeated testing situations that encourage learning or practice effects.

### 3.2 Demographics of participants in case control series

Given the maximal deviation in smell early in acute illness, we chose to explore the specific changes in multiple chemosensory modalities in the handful of cases who enrolled *prior* to onset of acute illness in greater detail. Demographics of four COVID-19 cases and four matched controls are summarized in Table 1. The mean age of participants with COVID-19 was 26 years of age (range 21-43) and the mean age of matched controls was 25.5 years of age (range 22-38).

### 3.3 Controls

Throughout the course of the study, controls exhibited normal function for smell, taste, and chemesthesis (Figure 3). Specifically, mean scores on odor identification, orthonasal smell intensity ratings, and ratings of perceived nasal blockage were relatively constant across days, although some participants were more variable than others. A few patterns deserve comment. For example, with Subject 1 we see a clear learning effect for odor identification where they became better at the task over time (Figure 3). Separately, controls may vary in perceived nasal blockage from day to day. For example, with Subject 3, we see a slight decrease in orthonasal smell intensity and a slight increase in nasal blockage in the last four days of testing (Figure 3). Such mild transient hyposmia would be wholly consistent with conductive smell loss due to nasal blockage typically seen with allergies or the common cold. Similarly, for taste, ratings of sweetness and sourness remained relatively constant throughout the course of the study for controls, although ratings were noisier for some participants than others. One rating for Subject 22 deserves comment: in the third week of testing, we observed a sharp drop in sour taste intensity and sharp increase in burn intensity, but only for a single day (Figure 3). While we cannot be sure, we suspect they simply picked the wrong cup from the bag of samples, as Sour Cherry jellybeans should not burn and should be sour. Subject 22 may have misread the 3-digit blinding code, or our research team may have mislabeled the cup. Either way, this single datum does not alter their overall pattern. Also, we note Subject 22 tends to give Cinnamon jellybeans a relatively high amount of oral burn relative to the other participants; we might speculate they eat spicy food infrequently, as large variation in burn ratings due to dietary exposure is very common (e.g., [44–47]).

**Figure 3:**
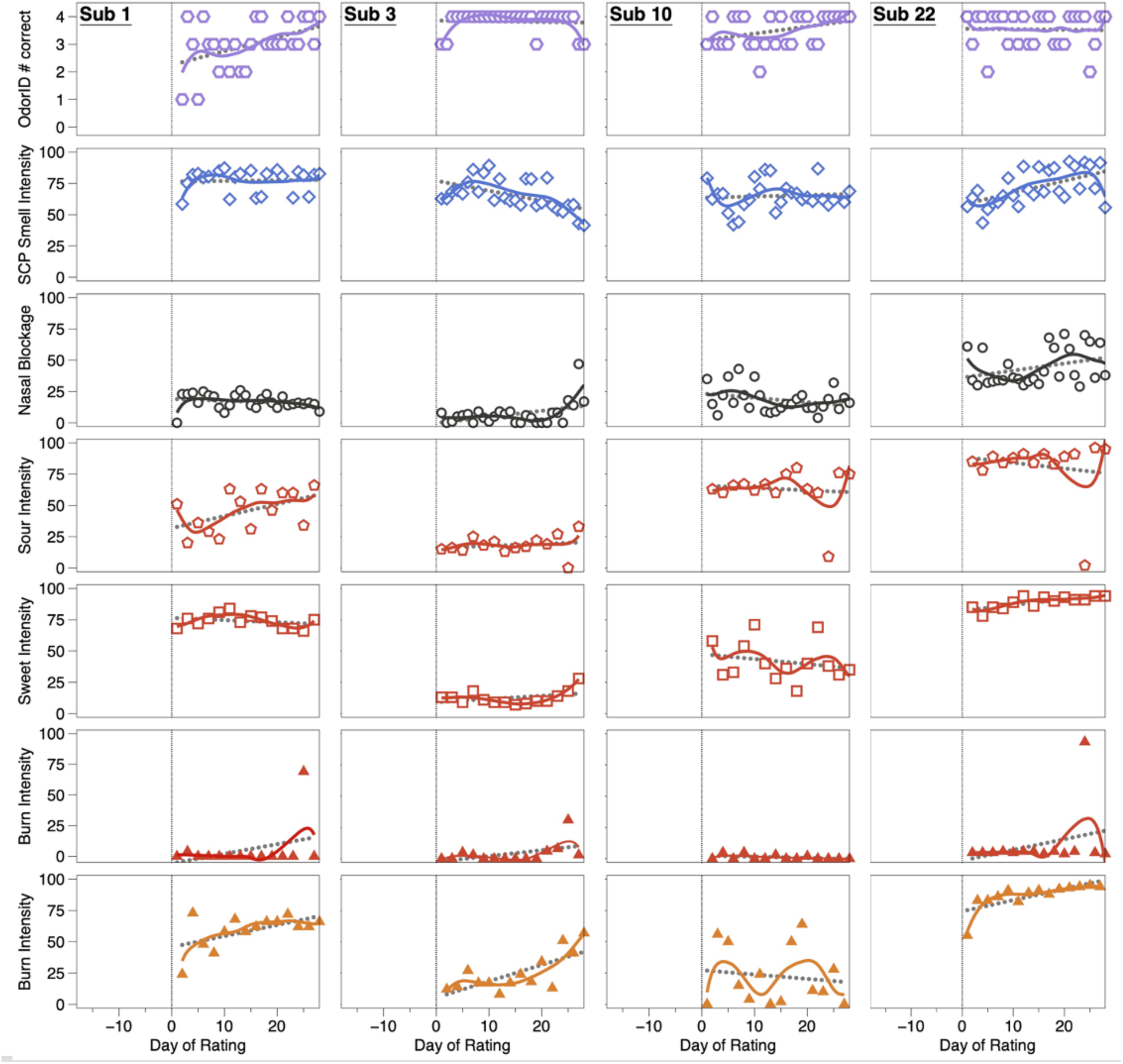
OdorID scores, and intensity ratings from matched controls over time. These participants (Subjects 1, 3, 10, and 22) show generally consistent ratings across the study. To help illustrate uniformity across the observation period, solid (colored) lines were fit via LOESS regression and dotted lines (gray) were fit via linear regression. A vertical line on Day 0 highlights the start of the 28-day study. Open hexagons (1^st^ row) are the number correct on a ScentCheckPro card, while open diamonds (2^nd^ row) are the mean daily smell intensity ratings from the same card. Open circles (3^rd^ row) reflect ratings of perceived nasal blockage. Red symbols (rows 4, 5, and 6) reflect specific quality ratings from Sour Cherry jellybeans collected with a pinched nose. Orange triangles (row 7) indicate burn ratings from Cinnamon jellybeans collected with a pinched nose.

In summary, daily data from these four *controls* suggest individuals without COVID-19 are able to correctly identify the odors from the ScentCheckPro cards, and to consistently rate the various attributes from the cards (orthonasal intensity) and the jellybeans (taste, burn). While some minor variation is observed over time and across participants, the ratings are generally stable over the study period, in sharp contrast to the COVID-19+ cases (Figure 4).

**Figure 4:**
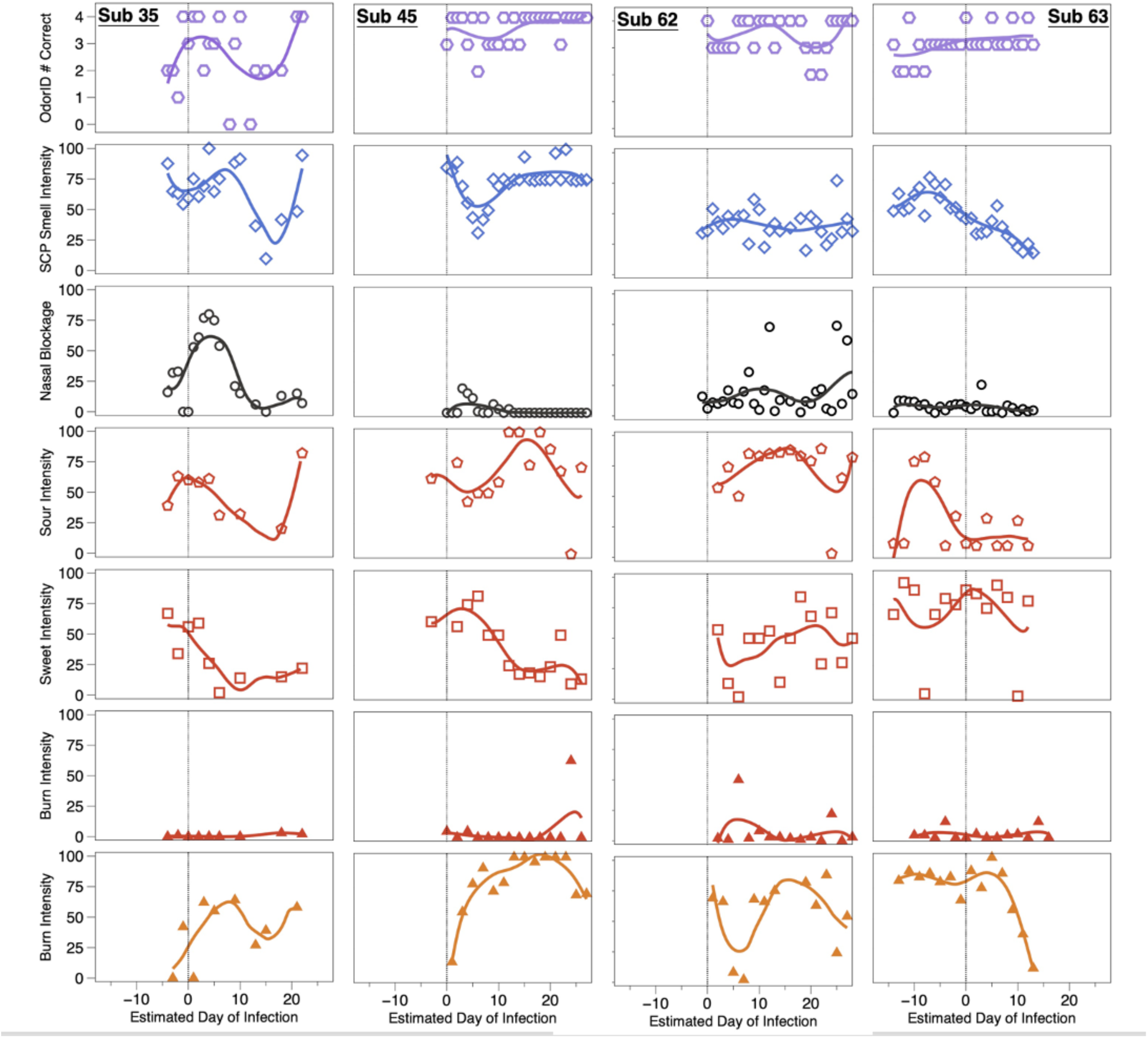
OdorID scores and VAS ratings from four COVID-19 positive individuals. COVID-19 cases (subjects 35, 45, 62, and 63) tended to show transient alterations of smell, taste, and/or chemesthesis during the observation period. To help illustrate uniformity across the study days, solid lines were fit via LOESS regression. A vertical line at Day 0 was added to highlight the estimated day of infection. Symbols and rows match those used in Figure 3: row 1 is daily number correct, and row 2 is orthonasal intensity of four scratch-n-sniff patches from a ScentCheckPro card, row 3 is ratings of perceived nasal blockage, rows 4-6 are sour, sweet, and burn ratings for Sour Cherry jellybeans with the nose pinched, and row 7 is burn for Cinnamon jellybeans with the nose pinched.

### 3.4 Cases

All four cases converted from being close contacts to being COVID-19+ while enrolled in the study, enabling visualization of changes in their responses over time (Figure 4). For brevity, we only highlight a few notable points here and a more detailed account is provided in the supplemental materials. When smell loss was observed, it was largely unrelated to nasal blockage, consistent with prior reports [4–7, 10] and the idea that COVID-19-associated smell loss arises from ACE2 receptor-mediated disruption of the olfactory epithelium, rather than the conductive losses typically seen with the common cold. Regarding chemesthesis, the lack of burn from the Sour Cherry jellybeans served as a negative control, suggesting participants were successful in discriminating between burn from a Cinnamon jellybean and a lack of burn from a Sour Cherry jellybean. From this, we can assume changes in burn observed here for the Cinnamon jellybeans was not merely a failure to understand the task.

#### 3.4.1 Subject 35

Symptoms for Subject 35 included cough, runny nose/congestion, sore throat, and headache. Odor intensity ratings dropped through day 15 before recovering; her OdorID scores showed a similar pattern, but her data were noisier given the learning effect noted previously. Her nasal blockage resolved around Day 8, but she still showed impaired smell. Sourness ratings declined until ∼Day 15 while sweetness declined until ∼Day 6, before each began to recover. This was not merely a taste/flavor semantic confusion, as ratings were obtained while wearing nose clips. Her data also suggest sweet and sour taste are each transiently affected with an active COVID-19 infection, and loss and recovery may be dyssynchronous (i.e., sweetness did not recover as swiftly as sourness). Separately, she showed large changes in burn from Cinnamon jellybeans, suggesting oral chemesthesis is affected by COVID-19 infection, and this may be dyssynchronous from altered taste or smell function.

#### 3.4.2 Subject 45

Subject 45 reported no symptoms despite becoming an active COVID-19 case while enrolled. Notably, despite being nominally asymptomatic, she clearly showed altered smell function that was reflected in both in OdorID performance and orthonasal intensity ratings, with maximal loss around Day 5. This highlights that some COVID-19+ individuals may be unaware of altered smell function, consistent with meta-analysis by Hannum and colleagues [8, 48]. As above, this transient disruption could not be attributed to nasal blockage. Regarding taste ratings, she also exhibited temporal dyssynchrony for different qualities. For the Cinnamon jellybeans, she showed a monotonic increase in burn over ∼3 weeks, before showing a small drop at the end of the study. Elsewhere, some patients reported an increase in the ability to feel sensations in the mouth (including burning) during recovery from COVID-19 [49], so her temporal pattern may potentially reflect acute hypoalgesia, followed by hyperalgesia, before eventually returning to normal. In any case, her data support the idea that oral chemesthesis can be affected acutely by SARS-CoV-2 infection.

#### 3.4.3 Subject 62

Like Subject 45, Subject 62 failed to report any symptoms, but unlike the prior cases, his orthonasal intensity ratings and OdorID performance remained relatively constant over time, and nasal blockage was generally low. This highlights that while many individuals with COVID-19 experience smell loss, some do not (e.g., [8, 48]). Regarding taste, noisy data preclude any strong conclusions, but tentatively, it seems he may have experienced large changes in both sweetness and sourness. That said, there is a sharp drop in sour taste intensity and sharp increase in burn intensity on two separate days (Figure 4). We suspect Subject 62 may have simply tasted the wrong sample on these days, as Sour Cherry jellybeans should be sour without any burn. Still, despite these caveats, his panel plots also suggest he experienced acute changes in oral chemesthesis without concomitant smell loss. If true, this would highlight that mechanisms of loss across all three chemosensory modalities are likely to be distinct.

#### 3.4.3 Subject 63

Subject 63 enrolled 2 weeks before becoming a case. This greatly exceeds the expected incubation period (5 to 7 days) [37, 50], so we contacted her via email and she reported a second exposure to a COVID-19+ individual. Thus, we assume she became ill upon her second exposure rather than the initial exposure that caused her to enroll. Her data reveal changes in smell, taste, and chemesthesis. However, the 28-day observation period only captures initial illness without recovery, as she initially enrolled after an exposure that did not cause infection. Consistent with this interpretation, she did not report any symptoms for the first 2 weeks, before reporting many symptoms (sore throat, fever or chills, dry cough, body aches, fatigue, diarrhea, nausea or vomiting, headache, and dry cough). Notably, her mean orthonasal ratings began to decline somewhat a few days before the estimated day of infection, in the absence of nasal blockage. Further, her intensity data suggest she experienced hyposmia, rather than full anosmia, so it is not surprising that her OdorID performance remained relatively constant across the study period. This suggests rated smell intensity might provide more nuanced assessment of smell function versus odor identification (as discussed above). Her taste data were somewhat noisy, but it seems sourness may have been more affected than sweetness. Tentatively, her plots suggest she lost taste function in a quality specific manner, along with partial smell loss and loss of oral chemesthesis, with staggered timing of each.

### 3.4 General Discussion

By using intensive longitudinal data collected daily for 28 days, we were able to assess COVID-19-associated changes over time. Further, by leveraging enrollment of individuals upon exposure rather than waiting until they were already ill, we were able to create a small case control series that captured both initial loss and recovery. We can draw several conclusions from the results described here. First, number correct on a short odor identification task may potentially miss dysfunction in hyposmic individuals who have clearly depressed perceived intensity, but who still retain enough function to successfully complete an identification task. Longer tests, such as the 40-question University of Pennsylvania Smell Identification Test (Sensonics), may be better at differentiating these levels of smell loss, though their greater cost and time-to-complete make them impractical for this type of intensive longitudinal study. Second, we find smell loss appears maximal around Day 5, but this varies somewhat across participants. Third, COVID-19 related chemosensory dysfunction can manifest as reduced oral chemesthesis, reduced taste, and/or reduced orthonasal smell with little to no nasal blockage, with temporally staggered onset and time course. Collectively, these results, although limited in scope, extend prior work by providing direct assessment of multiple sensory modalities repeatedly over time using stable, commercially-available products as stimuli.

A strength of this study involved the use of commercial stimuli like jellybeans to collect ratings while the nose was blocked with nose clips. First, because of their glazed candy shell, jellybeans have little to no orthonasal scent and the odorant is only released from the food matrix upon chewing. For our purpose, this gave us a convenience way to deliver consistent shelf stable stimuli safely during a pandemic. All jellybeans of the same flavor came from the same lot; given routine quality control measures in commercial manufacturing, we are confident participants received consistent stimuli even if we do not know the exact formulation of the jellybeans. Second, our use of similarly colored jellybeans with different flavors minimizes potential biases participants may have from prior experience or knowledge with other foods (e.g., this is a lemon, and I know lemons tend to be sour [51]). When this lack of expectation is coupled with the use of nose clips, we believe the data shown here reflect true differences in taste and chemesthesis and not merely they result of a flavor taste confusion. Other studies have also reported loss in function when using taste stimuli that do not have an olfactory component [52].

Currently, there is disagreement regarding whether different types of taste cells are differentially affected by SARS-CoV-2 infection. Indeed, several studies have showed quality-specific differences (i.e., [27, 43, 53–56]), while others have not (i.e., [18, 40–42, 57–61]). Here, sourness and sweetness from consistent stimuli were lost and recovered at similar rates for some participants, but this was not uniformly true. This implies specific taste qualities may recover at different rates, although additional work is needed to confirm this. Also, our data indicate taste qualities may be differentially affected, consistent with other reports [19, 62]. While the specific mechanisms underlying taste dysfunction with SARS-CoV-2 infection remain unclear, several mechanisms have be proposed. For example, ACE2 could allow for the infection of Type 2 (sweet, bitter and umami) taste receptor cells by SARS-CoV-2. Saliva could affect gustation as salivary glands express high levels of ACE2 and TMPRSS2. SARS-CoV-2 may even affect the central nervous system, as the virus has been detected in the cerebrospinal fluid [63–67].

Anecdotal reports and preliminary psychophysics suggest loss of chemesthesis with SARS-CoV-2 infection is real [10, 23, 30, 68]. We extend prior reports here by showing oral chemesthesis, not just nasal chemesthesis, may be altered by COVID-19. This effect appears to be highly transitory, which could cause underreporting, especially when assessment occurs multiple days after illness has started. Definitionally, chemesthesis includes both thermal and tactile percepts like warming, cooling, and buzzing, and these sensations occur via distinct and specialized receptors. Even if focusing solely on burn, multiple receptors like TRPV1 and TRPA1 are involved. Despite multiple advantages of commercial stimuli (high consistency, low cost, shelf stability, etc.), use of commercial jellybeans here limits interpretation somewhat. That is, cinnamon flavored candies presumably contain cinnamaldehyde, a well-known TRPA1 agonist [69, 70].However, we cannot rule out whether they contain capsaicin (or another TRPV1 agonist), as food labeling laws in the United States allow manufacturers to declare such ingredients as natural or artificial flavors on the package without being more specific, so we cannot make strong inferences about which specific chemesthetic mechanisms might be affected by SARS-CoV-2 infection. Nonetheless, present data extend prior work by clearly showing oral burn can be transiently affected by COVID-19.

## 4. Limitations and Conclusions

Our data suggest intensive cohort study designs are imperative for understanding and tracking symptoms of COVID-19 patients. Through intensive daily ratings we were able to examine and follow participants from initial exposure to catch symptoms as the emerged, allowing for the visualization of symptom onset, not just recovery. Thus, a strength of this study is the nature of the cohort examined, and it exemplifies a need for more cohort studies to catch patients before and during the most infectious period of their illness. A few limitations should be briefly noted. First, all sensory testing in this study was performed remotely at home, due to pandemic related safety restrictions meant to protect both participants and our research team. Because participants made daily ratings at their leisure without direct supervision, we cannot obtain the same level of stimulus control we would have with an in-person lab-based study. Also, while all study materials were clearly labeled, we cannot preclude whether participants may have occasionally chosen the incorrect blinding codes on some days or that our staff might have mislabeled these samples. Further, we should note the commercial ScentCheckPro scratch-n-sniff cards used here were not validated as a clinical smell test; also, they were originally designed as an odor identification task, rather than a smell intensity task, so we cannot assume all stimulus concentrations were precisely matched for intensity. This concern is partially offset however by the randomization of odorants on any given card, and the use of daily means. Finally, we fully acknowledge this study had a very small number of participants (albeit with many data points per participant), so present findings should be taken as tentative until confirmed. Attempts to generalize the incidence or prevalence of distinct types of loss or dysfunction should not be made from this small case-control series. Despite these limitations, this dataset is highly unique in that it captures changes very early in COVID-19 illness with intensive daily sampling.

Here, we extend current knowledge by showing oral chemesthesis, taste, and/or orthonasal smell function can each be acutely affected by COVID-19. Further, we find such disruption may be dyssynchronous for the different chemical senses, with differing rates of loss and recovery across modalities and individuals. Also, odor intensity ratings revealed potentially hyposmic individuals who might be missed if smell function is only assessed via odor identification scores. Finally, disrupted chemosensation, especially for chemesthesis, appears to be highly transient, suggesting studies that collect a single snapshot in time, often retrospectively, may underestimate the true prevalence of loss.

## Data Availability

All data produced in the present study will be made available upon reasonable request to the authors.

## 5. Acknowledgements

The authors wish to thank our participants for their time and critical contribution to this work.

## Supplemental Materials

**Supplemental Figure 1.**
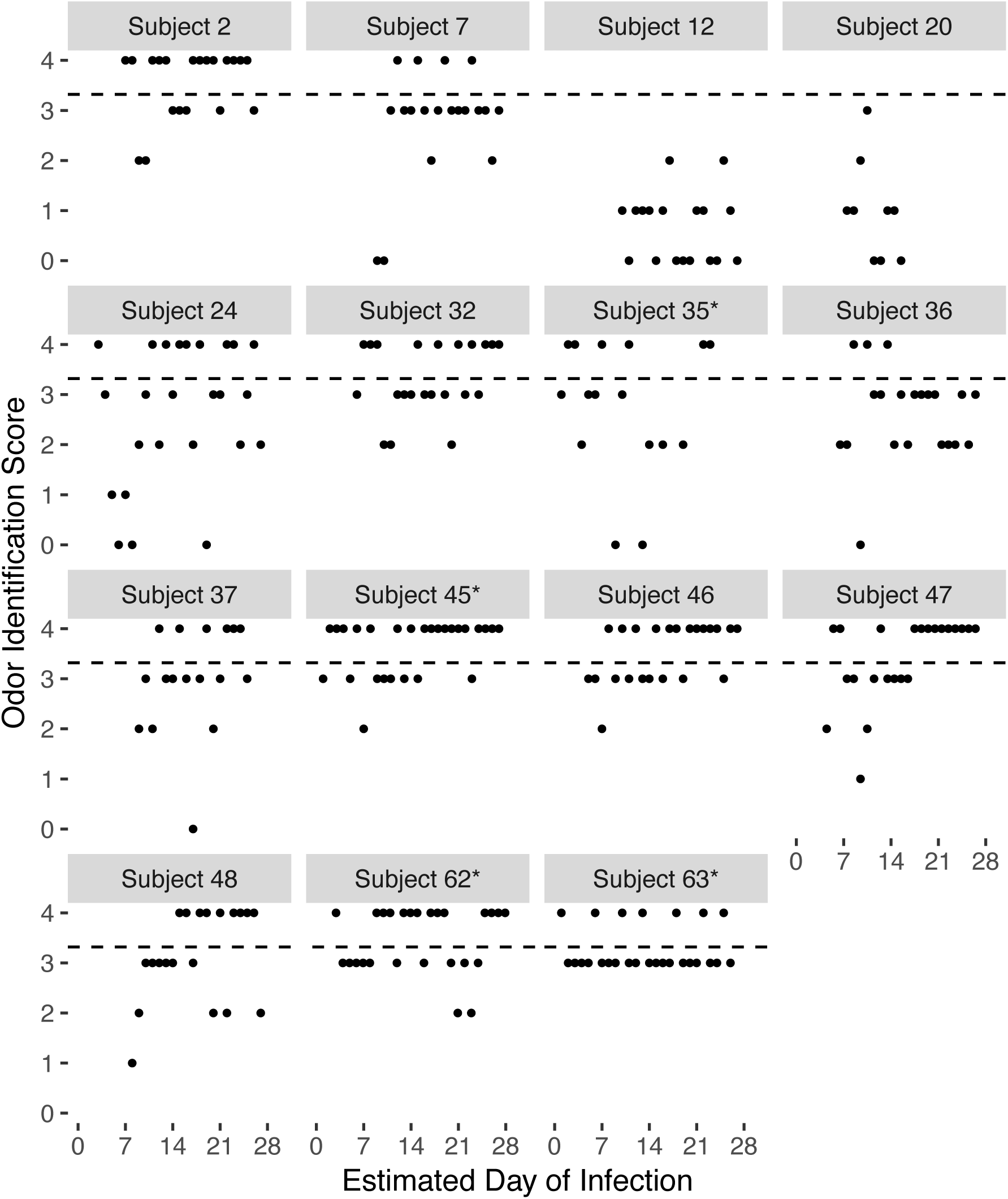
Raw daily odor identification scores from the ScentCheckPro cards over time for the COVID-19 cases (n=15) who entered the study on or before day 10 of their infection.

**Supplemental Figure 2.**
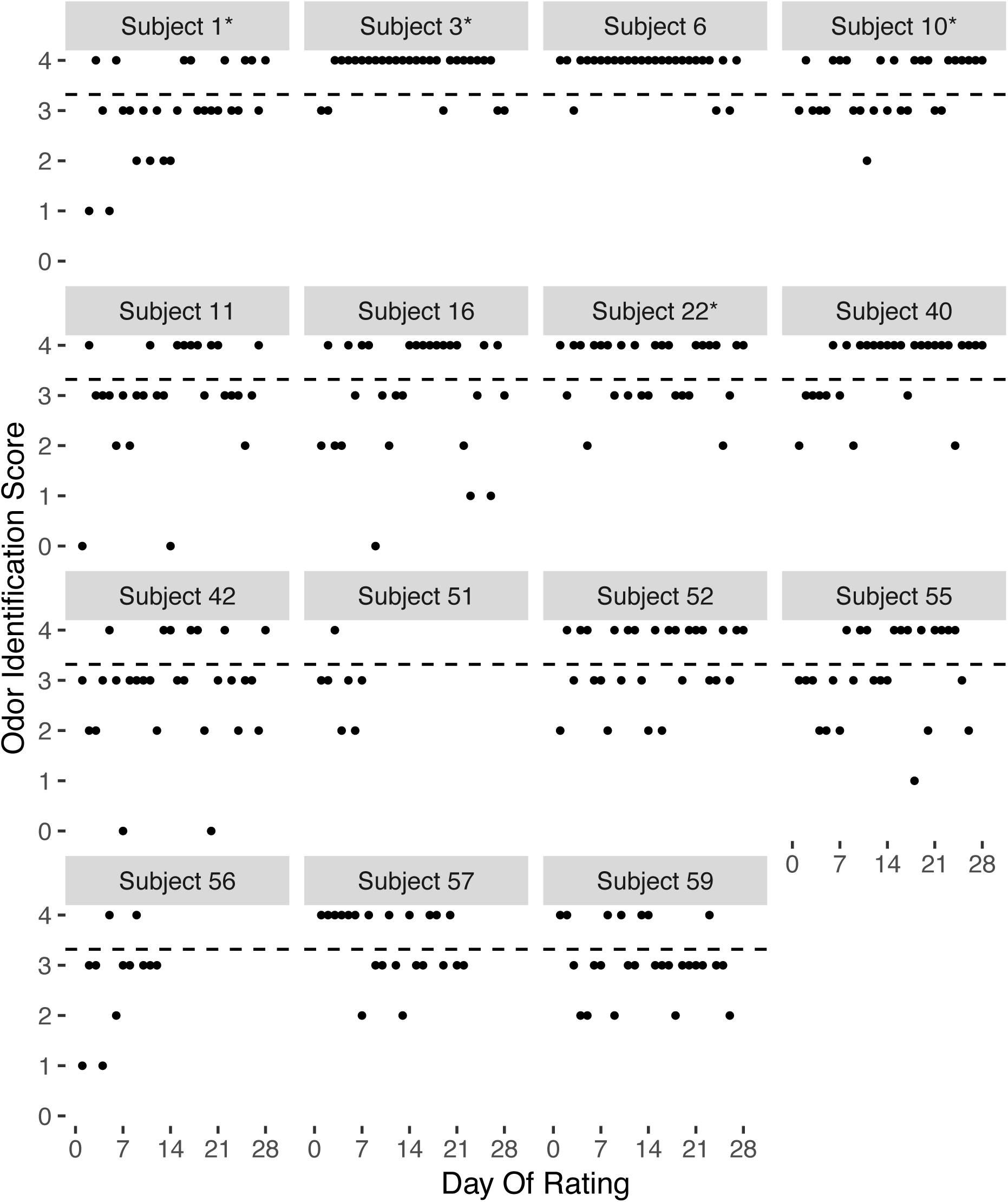
Raw daily odor identification scores from the ScentCheckPro cards over time for the 15 controls.

## Supplemental narrative and discussion of symptoms for the 4 cases

### Subject 35

Subject 35 became a case during the study, enabling visualization of the falling and rising phases of her responses (see Figure 4). Her symptoms included cough, runny nose/congestion, sore throat, and headache. Regarding orthonasal scratch-n-sniff intensity ratings, daily means decreased through day 15 before recovering. OdorID scores showed a similar pattern, but data were noisier given the learning effect described above. Together, this suggests her orthonasal olfaction was transiently affected during active COVID-19 infection, as expected (e.g., [19, 71, 72]). Notably, she showed impaired smell even after nasal blockage resolved around Day 8, and her maximal smell loss and maximal nasal blockage were dyssynchronous. This is consistent with other reports showing COVID-19 smell loss is not associated with nasal blockage [4–7, 10], presumably because COVID-19-associated loss arises from ACE2 receptor-mediated disruption of the olfactory epithelium, and not the conductive losses seen with the common cold.

Her sourness ratings from the Sour Cherry jellybean declined until ∼Day 15, when ratings began to increase, while sweetness declined until ∼Day 6, before beginning to recover. The decline and subsequent rise of sweet and sour taste likely signifies normal recovery, although Figure 4 also shows dyssynchronous recovery of these tastes (i.e., sweetness did not recover as swiftly as sourness). These data indicate sweet and sour taste are each transiently affected with an active COVID-19 infection, and this was *not* merely a taste/flavor semantic confusion, as ratings were obtained while wearing nose clips. Subject 35 also showed large changes in burn from the Cinnamon jellybeans, suggesting oral chemesthesis is affected by COVID-19. The lack of burn from the Sour Cherry jellybeans serves as a negative control, indicating she was successful in discriminating between burn from a Cinnamon jellybean and a lack of burn from a Sour Cherry jellybean (a pattern also seen in the three other cases shown in Figure 4). These data indicate perception of oral burn can be affected by an active COVID-19 infection dyssynchronously from taste or smell. While patient anecdotes (including social media posts) have previously suggested nasal and/or oral chemesthesis may be affected by SARS-CoV-2 infection [39, 49, 73], the daily assessment and prospective design used here provide quantitative evidence of altered oral chemesthesis with COVID-19.

### Subject 45

Subject 45 converted from being a close contact to an active COVID-19 case during the study, but unlike Subject 35, Subject 45 never reported any symptoms during her infection. Yet. despite being nominally asymptomatic, she still showed a clear drop in both OdorID performance and ratings of orthonasal intensity around Day 5 (with a bigger effect size for intensity). This highlights that some individuals infected with SARS-CoV-2 may be unaware of the impact on their sensory abilities, consistent with recent meta-analysis by Hannum and colleagues [8, 48]. Nor was this transient disruption in smell due to nasal blockage (as reported elsewhere [4–7, 10]). As her infection progressed, sourness from the Sour Cherry jellybean was variable, and sweetness from this jellybean steadily declined over the course of infection, again indicative of temporal dyssynchrony for different taste qualities. In contrast to burn rating from the Sour Cherry jellybean (which stayed near 0 across the study period, as expected), burn from the Cinnamon jellybean steadily increased in a monotonic fashion until a small drop was observed at the end of the study. Taken together with data from Subject 35, this indicates suggests oral chemesthesis is altered by active SARS-CoV-2 infection. Another study noted that during recovery from COVID-19, some patients report an increase in the ability to feel sensations in the mouth, including burning [49].

### Subject 62

Subject 62’s infection began 1 day before enrollment. Like Subject 45, he failed to self-report any symptoms, but unlike Subjects 35 and 45, his orthonasal intensity ratings and OdorID performance remained relatively constant throughout the study period, and his nasal blockage was generally low – while many individuals experience smell loss with COVID-19, some do not (e.g., [8, 48]). Regarding taste, noisy data make it hard to draw any strong conclusions, but it still seems he may have experienced substantial changes in sour and sweet taste. Regarding burn, he rated the burn from Sour Cherry jellybeans near zero for the entire study, suggesting he successfully distinguished burn from the Cinnamon jellybean from the lack of burn from the Sour Cherry jellybean (like the other cases). Two other values merit comment: in the 1^st^ and 3^rd^ week of testing, a sharp drop in sour taste intensity and sharp increase in burn intensity can be seen on two separate days; we suspect he may have misread the blinding codes, tasting the wrong sample on these days, as Sour Cherry jellybeans should be sour without any burn. Still, despite noise in his ratings, his panel plots for burn suggest he experienced transient changes in oral chemesthesis. If this case did in fact experience altered burn and altered taste without concomitant smell loss, this would emphasize that mechanisms of loss across all three modalities are distinct, with the caveat that the noise in these data should temper any strong inferences.

### Subject 63

Subject 63 enrolled 2 weeks days before becoming a case. Because this greatly exceeds the expected incubation period of 5 to 7 days [37, 50], our study team contacted her via email. At that point, she reported a second exposure to an individual with COVID-19 – we assume this second exposure was the source of the infection documented here. Her data reveals changes in smell, taste, and chemesthesis as she transitioned from being a close contact to being a case, but the observation period only captures her initial illness without any recovery as she had enrolled after her first exposure that did not cause an infection. Consistent with this interpretation, she did not report any symptoms for the first 2 weeks, but then began reporting many symptoms (sore throat, fever or chills, dry cough, body aches, fatigue, diarrhea, nausea or vomiting, headache, and dry cough). Notably, her mean orthonasal ratings began to decline somewhat a few days before the estimated day of infection, but she indicated little to no nasal blockage, as expected [4–7, 10]. Also, her intensity ratings suggest she experienced hyposmia, rather than full anosmia, so it is unsurprising that her OdorID performance remained relatively constant across the study period, with some evidence of a slight learning effect near the beginning of the study. As discussed previously, this suggests rated smell intensity might provide more nuanced assessment of smell function versus odor identification. We have no obvious explanation for her unexpectedly low sourness ratings on the first two days of the study. Still, if her peak ratings during this initial (uninfected) period are tentatively treated as a baseline, we see a subsequent decline in sourness around the time her other symptoms appeared. For the rest of the study, her sour ratings remained relatively depressed, at least relative to the maximal values she reported pre-infection. In contrast, sweetness, while noisy, appeared more constant across the entire study. Tentatively, these plots suggest Subject 63 lost some taste function in a quality specific manner, as well as partial smell loss and loss of oral chemesthesis, with staggered timing of each, during her infection.

